# Learning Decision Thresholds for Risk-Stratification Models from Aggregate Clinician Behavior

**DOI:** 10.1101/2021.02.19.21252069

**Authors:** Birju S. Patel, Ethan Steinberg, Stephen R. Pfohl, Nigam H. Shah

## Abstract

Using a risk-stratification model to guide clinical practice often requires the choice of a cutoff - called the decision threshold - on the model’s output to trigger a subsequent action such as an electronic alert. Choosing this cutoff is not always straightforward. We propose a flexible approach that leverages the collective information in treatment decisions made in real life to learn reference decision thresholds from physician practice. Using the example of prescribing a statin for primary prevention of cardiovascular disease based on 10-year risk calculated by the 2013 Pooled Cohort Equations, we demonstrate the feasibility of using real world data to learn the implicit decision threshold that reflects existing physician behavior. Learning a decision threshold in this manner allows for evaluation of a proposed operating point against the threshold reflective of the community standard of care. Furthermore, this approach can be used to monitor and audit model-guided clinical decision-making following model deployment.

## INTRODUCTION

Physician informaticists are increasingly involved in the deployment of risk-stratification models for clinical decision support. They may be asked if the predictive performance, such as the sensitivity and specificity, of a machine learning-derived model is acceptable for guiding the allocation of an intervention in their health system.[1] Given this information, a corresponding operating point on the receiver operating characteristic (ROC) curve is used as a cutoff for an action trigger, such as generating an alert for the early detection and treatment of sepsis.[2]

Medical decision analysis, one of the most recognized approaches for determining an operating point, requires economic utility values for correct and incorrect predictions as inputs to calculate a decision threshold.[3,4] However, these inputs can be difficult to obtain in practice.[5,6] Alternatively, it is possible to obtain a decision threshold by having physicians respond to a series of clinical vignettes where risk scores are known.[7–11]

We recognized an opportunity to augment such evaluation beyond hypothetical clinical cases using real data from the collective practice of many physicians recorded by the electronic health record.[12] We hypothesized that clinician behavior—reflecting how physicians across an organization balanced harms, benefits, costs, patient preferences, and resource constraints to make shared clinical decisions with actual patients in individual situations[13]—could be used to learn the latent decision threshold that was used in practice.[14] This learned threshold could then be used as a reference[15] to understand how a potential operating point compares to the current community standard of practice when deploying or monitoring a risk-stratification model. Our objective was to demonstrate the feasibility of a flexible mathematical approach that uses observational data to learn the underlying decision threshold implicit in physician practice. To illustrate the approach with a clinical example, we fit an equation that best captured clinical decision-making from observational data, extracted decision thresholds from this equation, compared these empirical results to guideline recommended thresholds, and assessed the stability of learned decision thresholds after the release of updated clinical guidelines.

## METHODS

As an example, we learned decision thresholds for statin treatment based on the 2013 Pooled Cohort Equations (PCEs),[16] which predict 10-year atherosclerotic disease risk and have well described decision thresholds.[17] We constructed a retrospective cohort from the Stanford Medicine Research Repository of adult primary prevention patients who underwent lipid screening by a primary care provider before 2013 and met criteria to have their 10-year risk of atherosclerotic disease calculated. We ascertained whether patients were prescribed a statin within 180 days after lipid screening and if they developed major atherosclerotic disease within the following 10 years.

Our method begins by developing a mathematical equation that fits the decision to prescribe a statin using the 10-year risk of atherosclerotic disease provided by the PCEs (Figure 1). The formulation of the equation is inspired by expected utility theory, where a decision to treat is made if the net utility from treatment is greater than zero.[18] For example, a provider will likely choose to prescribe a statin if the decreased risk of atherosclerotic disease outweighs the potential side effects of the medication. We then specify net utility as a function of disease probability,[19] which is the output of the risk-stratification model, and use a sigmoid function to link increasing net utility to increasing probability of treatment. This produces the general form of the decision-making equation:

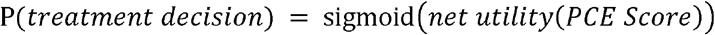

**Figure 1.**
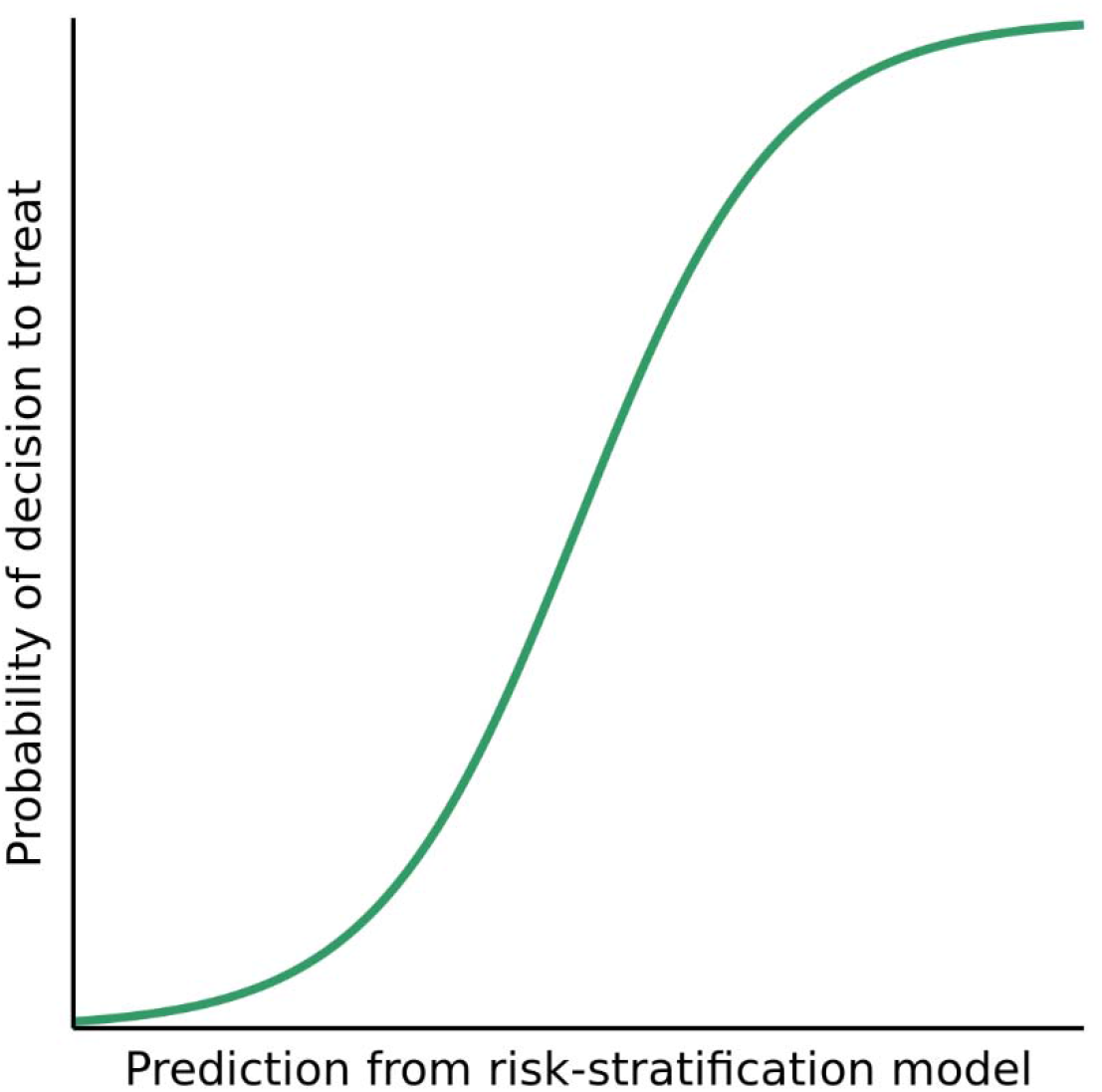
Hypothetical example of a decision-making equation. A decision-making equation (green line) predicts the probability of observing a decision to treat, shown here on the y-axis, as a function of the predictions from a risk-stratification model, shown here on the x-axis.

We fit two alternative equations from this general form using real-world data. In the first, we define net utility using a linear transformation of the PCE risk score and specify the standard logistic function as the choice of sigmoid function:

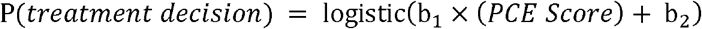

where b_1_ and b_2_ are coefficients that are learned from the data. In the second form of the equation, following observations on the impact of risk tolerance on utility,[20,21] we define net utility using a logarithmic transformation of the PCE risk score:

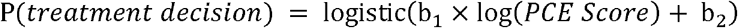

We evaluate which equation best captures real world decision-making as evidenced by the lowest Brier score, a measure of model fit. This decision-making equation is used to identify the decision thresholds as follows: for a specified predicted probability of treatment, we use the equation coefficients to solve for the corresponding risk score. We examine two specific treatment probabilities. The first, where the probability of treatment is equal to the overall treatment proportion in the cohort, reflects the risk score where clinicians commonly prescribe statins. We refer to this threshold as the *popular vote*. The second, where probability of treatment is equal to fifty percent,[8,14] reflects the risk score where net benefits become greater than harms. Since the equation predicts that more than half of patients with risk scores at or above this point are treated with statins, we refer to this threshold as the *majority vote*. We then compare these empirically derived thresholds with thresholds stated in guidelines.

Finally, to evaluate the sensitivity of these results to the release of updated clinical guidelines in 2013, we generate a cohort of patients screened for ASCVD risk after 2013 and examine whether there are differences in the equation fit or derived thresholds.

## RESULTS

Of the 4,705 patients seen at Stanford Medicine between 2009 and 2013 who underwent primary prevention risk assessment, 1,045 (22.2%) were prescribed a statin. The PCEs had similar discriminative ability (c-statistic 0.71) in this cohort compared to the original cohorts in which they were constructed.[16] The median 10-year risk score calculated by the PCEs was 2.4% in those not treated and 6.1% in those treated with statins.

As expected, we found that increasing 10-year atherosclerotic disease risk was associated with higher rates of prescribing statins (Figure 2). The log transformed equation better fit observed clinician decision-making than the linear one (Brier score 0.159 for the log transformation vs 0.165 for the linear transformation). In the log transformed equation, the average treatment rate (22.2%) corresponded to a popular vote decision threshold of 3.6% 10-year risk. The fifty percent probability of treatment corresponded to a majority vote decision threshold of 23.0% 10-year risk.

**Figure 2.**
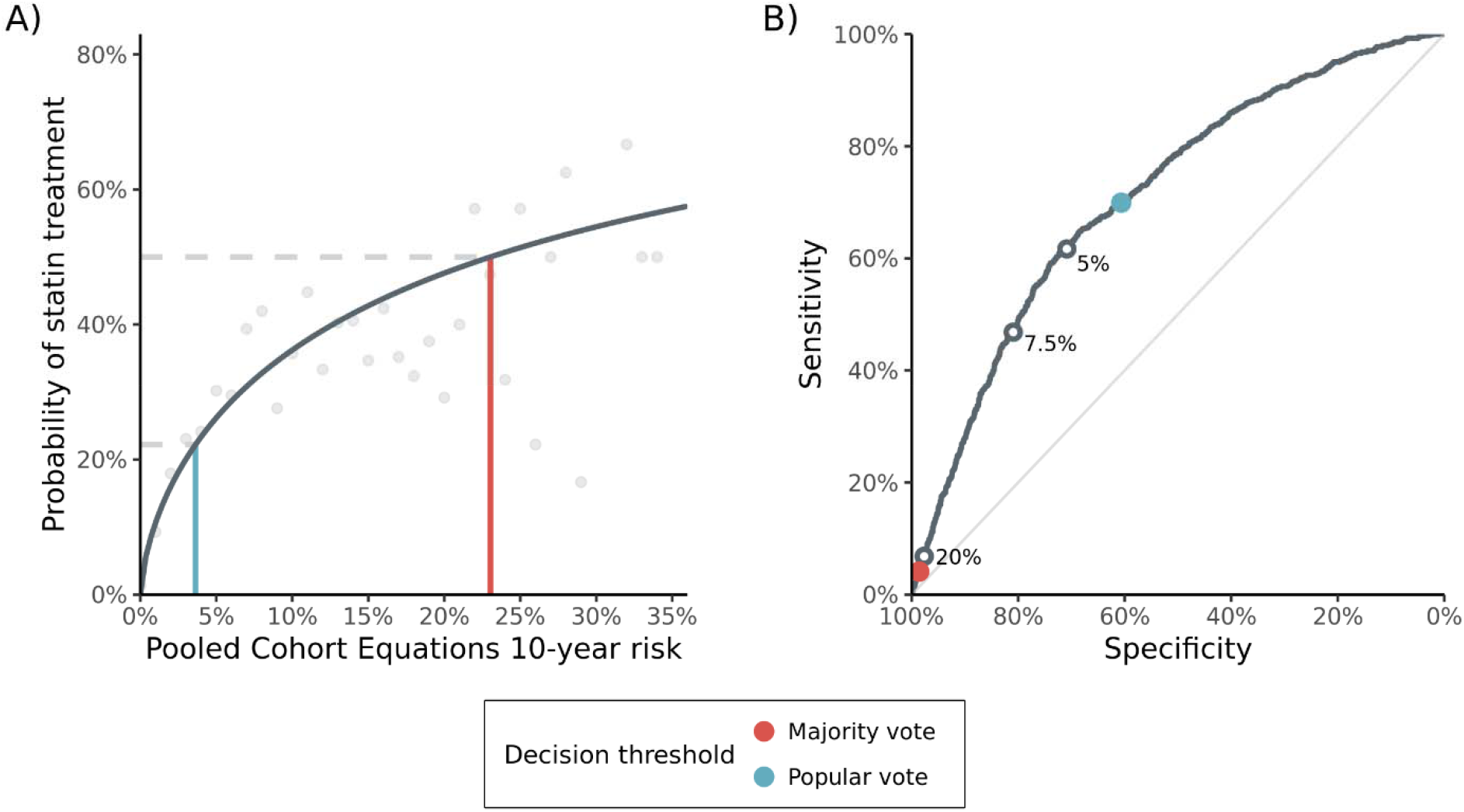
Decision Thresholds Derived from the Decision-Making Equation. A) The fitted decision-making equation (dark grey line) captures the relationship between the risk score and the decision to prescribe a statin (light grey circles are treatment rates for patients binned to the nearest whole number risk score percent). The cohort average treatment rate with a statin (lower dashed line) corresponds to the popular vote decision threshold of 3.6% 10-year risk (blue line), while the fifty percent probability of statin treatment (upper dashed line) corresponds to the majority vote decision threshold of 23.0% 10-year risk (red line). B) The ROC curve demonstrates the relationship between sensitivity and specificity at various potential thresholds. To make decisions based on the output of the PCEs, a continuous risk score is converted to a dichotomous recommendation by setting an operating point on the ROC curve. Published clinical guidelines set operating points at 5%, 7.5%, and 20% 10-year risk (white circles), which are located near the popular vote (blue circle) and majority vote (red circle) operating points learned from the decision-making equation.

The PCEs are essentially a predictive model whose continuous risk score is converted to a treatment recommendation by setting an operating point on its ROC curve (Figure 2). Based on the observed decision-making behavior from 2009 to 2013, an operating point set near the popular vote decision threshold would capture patients in the borderline (5%) and intermediate (7.5%) risk categories of the 2013 guidelines. An operating point set near the majority vote decision threshold would capture patients in the high-risk category, where the guideline uses a cutoff of 20% and documents the large benefits associated with high-intensity statin therapy for these patients.

We then constructed a cohort of patients who had risk assessments performed after the publication of the PCEs in 2013. This cohort included 23,291 patients, of whom 4,851 (20.8%) were treated with statins. We found a similar pattern of decision-making (Figure 3) after the guidelines were released, with the recalculated decision-making equation having similar performance (Brier score 0.148). The popular vote decision threshold was 3.9% and the majority vote decision threshold was 23.7%, which were 0.3% and 0.7% higher than the thresholds from the pre-2013 cohort, respectively.

**Figure 3.**
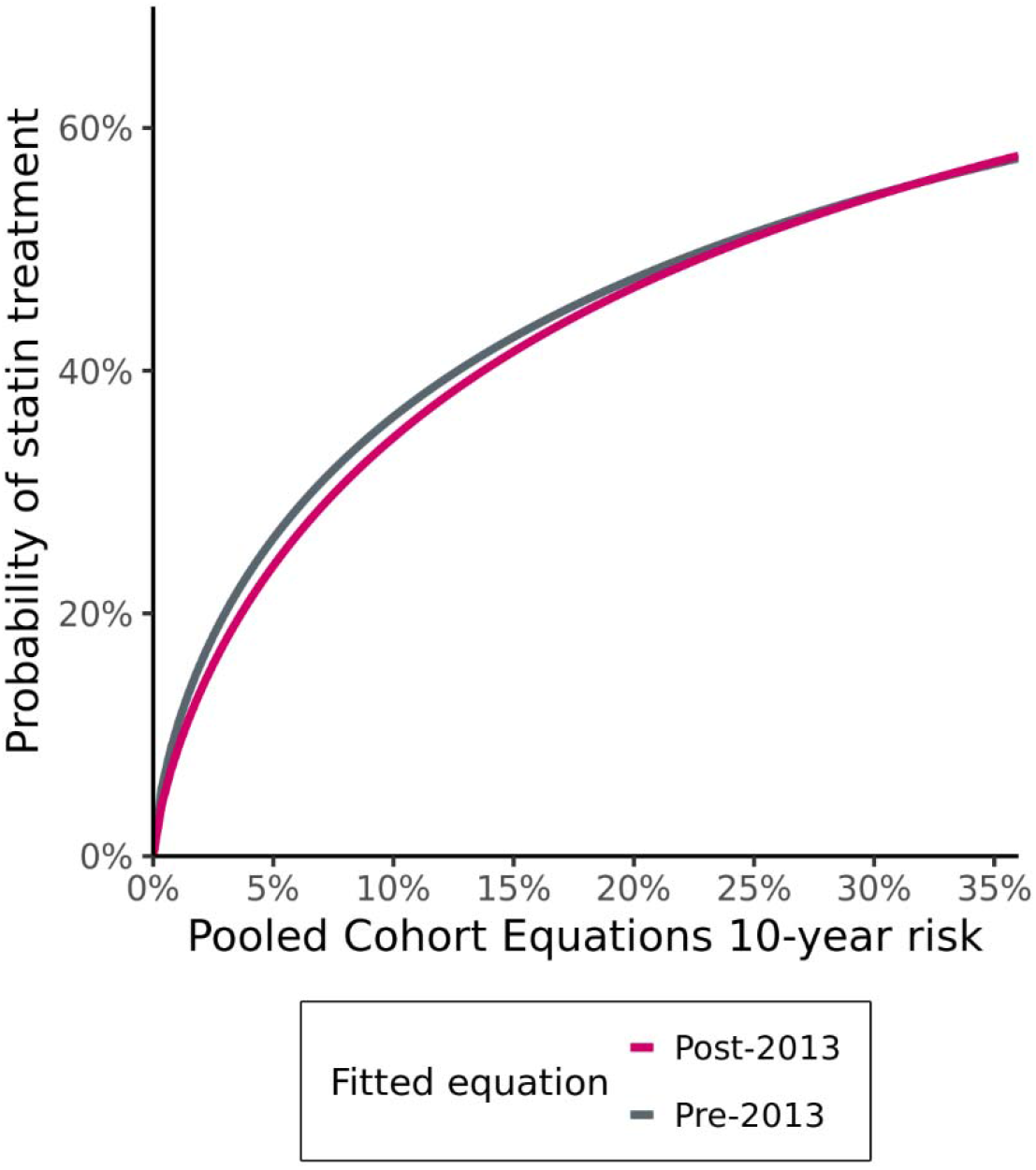
Comparison of Fitted Decision-Making Equations for the Pre- and Post-2013 Cohorts. The fitted decision-making equation for the pre-2013 (grey line) and post-2013 cohorts (magenta line) closely overlap over the range of observed PCE risk scores. This results in similar derived decision thresholds before and after publication of updated clinical guidelines in 2013.

## DISCUSSION

We find that an analysis of past, aggregate physician behavior can translate the combination of revealed patient preferences, clinical judgement, and decision rules used in practice into reference operating points on the ROC curve of a risk prediction model. To our knowledge, this is the first example describing how empirical decision thresholds learned from past clinician behavior of ordering an intervention could inform the deployment and monitoring of a risk-stratification model. For the PCEs, our data-derived thresholds demonstrate how collective physician behavior concurred with the decision thresholds later determined by an expert guideline panel. In addition, the popular vote decision threshold we found for statin prescribing was located near the decision threshold suggested by a well-done cost-effectiveness analysis,[22] providing reassurance that our approach captured similar information from real life decision-making as decision analysis does when utility values are known.

The cholesterol treatment guidelines in effect before the PCEs[23] used a strategy based on tiered cholesterol targets to recommend medical therapy. When the PCEs were released with a new risk-based framework in 2013, our method could have been used to translate the existing community standard of practice into reference decision thresholds for the new risk-stratification model. Such context could have been useful to communicate with physicians who were weary of how to reconcile these new risk scores with their practice.[24] One particular benefit of the approach we describe is that it can be used even when a risk-stratification algorithm does not already exist. With the proliferation of risk-stratification models in healthcare,[25] there will be many novel use cases where understanding current clinical practice will be relevant for model implementers.

The proposed method can also be used to monitor patterns of decision-making after the deployment of a risk-stratification model to identify changes and biases in clinician behavior. For example, we observed only minor changes in learned decision thresholds after 2013, suggesting physician behavior in practice was similar even when a new algorithm to calculate cardiovascular risk was recommended. The method could also be used to assess how decision-making differs across relevant subgroups while controlling for the risk of the outcome.[11] Observing a difference in the learned thresholds may be evidence of a differential standard of care and could generate hypotheses for further auditing to identify disparities in care processes and model derivation.[26] The method could also be extended to evaluate differences in decision-making patterns among individual physicians.[7,14,27]

We derived two different decision thresholds from the decision-making equation. It may be helpful to consider different thresholds when intending to use a risk-stratification model to reduce errors either due to omissions or commissions of care.[28] For example, an operating point near a popular vote decision threshold could flag deviances in care when patients with risk scores lower than that threshold are prescribed interventions, exposing them to potential side effects and unnecessary costs. On the other hand, an operating point near the majority vote threshold could alert a provider who has not yet prescribed a high-risk patient a potentially beneficial treatment, avoiding an omission in care. In contrast, setting a single decision threshold between these ranges attempting to reduce both omission and commission errors may trigger excessive alarms, resulting in overridden alerts[29] and provider fatigue.[30]

Our approach differs from other methods to evaluate the choice of decision thresholds by permitting flexibility for the multiple components of decision-making that are not often explicitly measured, such as patient risk tolerance and individualized estimates of harms and benefits. In contrast, medical decision analysis requires an upfront measurement of utility values to complete the decision modeling process.[3,31] A different approach, decision curve analysis,[5] is useful to highlight the range of decision thresholds where the model contributes predictive value but is not able to determine on its own if a clinician or patient would think those thresholds are reasonable given real-world context, which our method directly observes.

One important contribution of our method is the flexibility in the choice of mathematical equation to fit treatment decisions. While prior efforts to learn decision thresholds have used linear transformations of risk scores,[8,14,32] we found that the logarithmic transformation performed better for statin prescribing. Clinical situations with fundamentally different consequences for harms and benefits may require even different non-linear transformations. For example, prospect theory suggests that utilities can have an exponential form and that harms may be weighed more heavily than benefits by decision-makers in real life.[33] Additionally, we used the logistic function in our equations to link net utility to the decision to treat, but other sigmoid functions are available. The approach we have described can accommodate any assumption that expresses net utility as a function of predicted risk, and an implementer may consider using the equation that best fits decision-making measured by an objective scoring function such as the Brier score.

Overall, learned decision thresholds can provide useful empirical information to evaluate the context of a potential operating point when using a risk-stratification model to guide care.

## Data Availability

The data that support the findings of this study are available on reasonable request from the corresponding author. The data are not publicly available due to information that could compromise the privacy of research participants.

## Author contributions

Dr. Patel had full access to all the data in the study and takes responsibility for the integrity of the data and the accuracy of the data analysis.

## Concept and design

Patel

## Acquisition, analysis, or interpretation of data

All authors

## Drafting of the manuscript

Patel

## Critical revision of the manuscript for important intellectual content

All authors

## Statistical analysis

Patel

## Administrative, technical, or material support

Shah

## Supervision

Shah

## Data sharing statement

The data that support the findings of this study are available from the corresponding author upon reasonable request. The data are not publicly available due to information that could compromise research participant privacy.

